# A Qualitative Study of the Ethical Issues Encountered at End-of-life Care at a University Teaching Hospital in Nigeria

**DOI:** 10.1101/2024.04.22.24305718

**Authors:** Nnenma Ndidiamaka Udeh, Idemili-Arounu Ngozi, Emmanuel R Ezeome

## Abstract

**Background:** End-of-life (EOL) care involves providing quality medical attention to the dying patient. It is fraught with some ethical challenges, often under-explored in African settings. This communication presents a qualitative analysis of ethical issues encountered by caregivers and their patients who are receiving end-of-life care in a teaching hospital in Nigeria.

**Methods:** Ethical issues in EOL care encountered by 40 people (dying patients, their families, nurses, and doctors) at the University of Nigeria Teaching Hospital, Enugu (UNTH-E) Enugu State, Nigeria was explored over two months. The Participants’ socio-demographic data was analyzed using descriptive statistics, while qualitative data was analyzed using a thematic framework. The coding of transcripts was done with NVivo 12 software.

**Results:** All participant groups encountered commonly reported ethical challenges in end-of-life (EOL) care, including issues of medical futility, treatment refusal, truthful disclosures, families requesting that a competent patient not be informed about their condition, confidentiality, limiting or withdrawing a treatment, limited or insufficient pain management, conflicting interests in care, an unfair financial burden without the patient consent, and an unfair burden on the healthcare system. Additionally, the uncommon issues included the patient’s unwillingness to discussions about their terminal status; families withdrawing due care and support prematurely, and delayed referrals.

**Conclusion:** Ethical issues are commonly encountered in caring for the patient at EOL in the Nigerian environment notwithstanding the paucity of literature on them. This underscores the importance of adopting known preventive measures to eliminate or minimize these issues.

## INTRODUCTION

End-of-life (EOL) care is the support provided to dying patients when life-prolonging treatment or control of the disease has stopped,^[1]^ ensuring they receive the best possible care while respecting their autonomy.^[2]^ Ethical challenges sometimes arise in EOL care,^[3][4]^ particularly with the increasing use of new medical technologies and advances.^[3][5]^

Autonomy issues are prevalent in American populations,^[6]^ arising from communication gaps, differing opinions, and fair treatment.^[7]^ Other reported issues were truthful disclosure, conflicting obligations, and futility.^[8]^ In Italy,^[9]^ treatment disagreements and futility were common, while in India,^[10]^ paternalistic approaches were common. In Iran,^[11]^ families requested that patient information be undisclosed. Most studies on EOL care are from Western countries or cultures, with scant information from Africa and Nigeria.^[13][14]^ A systematic review in sub-Saharan Africa identified poor information communication by health personnel in South Africa and Uganda,^[12]^ and ethical case deliberation in Nigeria highlighted issues concerning patient autonomy, informed consent, communication gaps, and collaboration.^[4]^

It is uncertain if these challenges do not exist or are unnoticed in our setting, but information on their existence and nature could guide practices, policies,^[15]^ and research among people receiving EOL care. In addition, the existing literature on ethical issues at the EOL lacks harmonized terminologies and a globally accepted classification system. We note that bioethical principles like respect for a patient’s autonomy, beneficence and non-maleficence, and justice often contribute to these ethical issues.^[16][17]^

In this study, we report on ethical issues in EOL care encountered at the University of Nigeria Teaching Hospital in Enugu (UNTH-E), Nigeria, and classify them based on the bioethical principles that generated them.

## METHODS

This cross-sectional qualitative study examines ethical issues faced by dying patients, their families, and health caregivers at the UNTH-E Oncology Center and intensive-care unit (ICU). The aim is to classify and provide empirical evidence on known and unique ethical issues in EOL care in this region. The research took place between March and May 2022.

UNTH-E is a renowned oncology center in Nigeria, offering comprehensive cancer care. It operates an outpatient clinic with a chemotherapeutic unit, an inpatient ward, and a functional palliative care unit. It is located in Enugu State and serves 21.9 million people in five south-eastern states,^[18]^ and neighboring states in Nigeria, particularly in oncology and intensive care.

The UNTH-E Health Research Ethics Committee granted ethical approval for the study, which followed the Nigerian National Code of Health Research Ethics and the Helsinki Declaration. The participants provided written informed consent before enrollment.

The study commenced with a comprehensive literature review on ethical issues in EOL care and categorized them into a coherent grouping system based on the dominant bioethics principles that generated them, including those from conflicts of bioethical principles. See Figure 1. This classification enabled a comprehensive identification of these ethical issues and formed the basis for empirical exploration in the study population.

**Figure 1:**
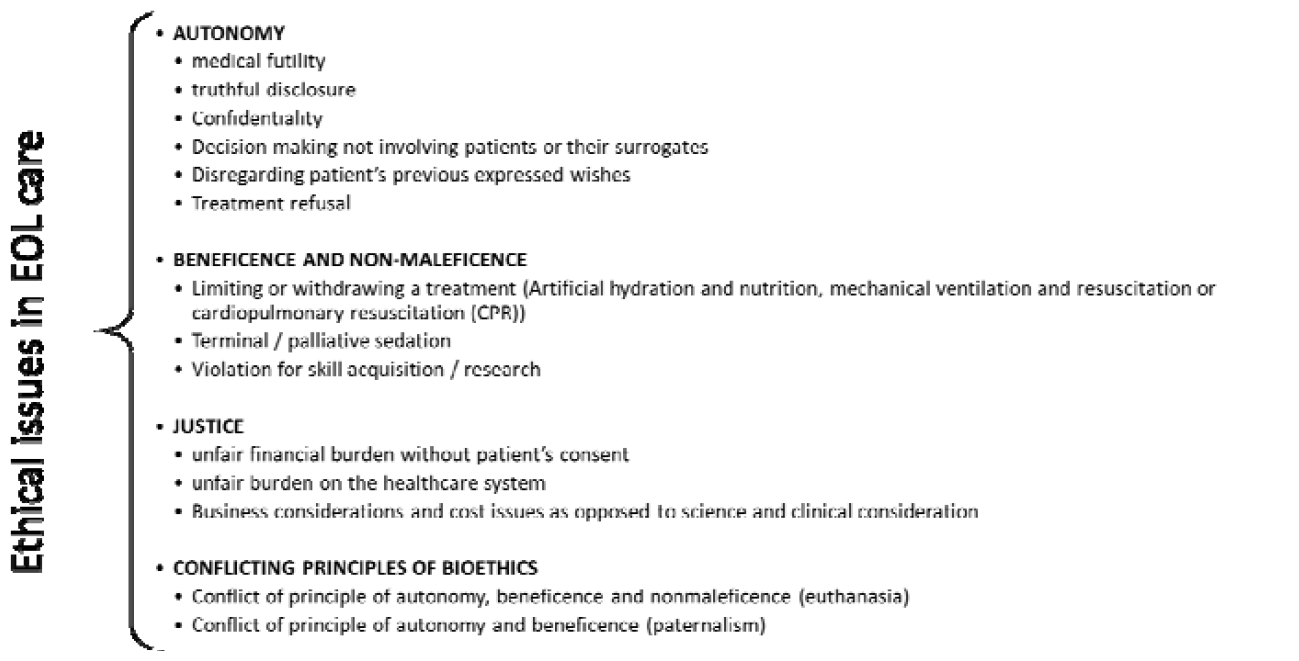
Ethical issues in EOL care

A purposive sampling method was employed to include adult patients in UNTH-E oncology wards and ICU receiving EOL care during the study period, their families (one per patient), and healthcare givers (senior doctors and nurses, one each). Exclusions encompassed patients experiencing weakness, pain, distress, isolation, family members, and health caregivers not involved in care for at least one week. Eligible participants were identified by the researcher, assisted by the nurse-in-charge of the oncology ward or ICU unit. After explaining the study, they were encouraged to consent and participate.

Typically, data saturation occurs after conducting 10–12 in-depth interviews within a homogeneous population.^[19][20][21]^ Therefore, a minimum sample size of ten patients, ten family members, and ten nurses and doctors each. Two data collection instruments were employed: a questionnaire/key-informant interview (KII) guide for nurses and doctors and a questionnaire/in-depth interview (IDI) guide for patients and their relatives. The questionnaires collected socio-demographic information and details regarding experience in ICU or oncology EOL care, post-qualification ethics training (for KII participants), or their duration of stay in the ICU or oncology-care wards (for IDI participants). Interview guides were developed based on current literature and pre-tested with four participants (a patient, their relative, a nurse, and a doctor) before use. A pilot study confirmed their effectiveness in prompting discussion and eliciting details on study objectives during interviews.

Participants were interviewed in UNTH-E wards and offices in the English language for 30 to 45 minutes. Interviews were audio-taped, transcribed verbatim, and supplemented with field notes.

The data was analyzed using descriptive statistics and a thematic framework, with transcripts coded in NVivo 12 by two independent individuals.

## RESULTS

### Sociodemographic

The study involved 20 KII participants aged 40–49 with 11–15 years of experience in oncology and ICU care, with 8 having post-qualification formal ethics training in workshops or as part of postgraduate training. Most IDI patients had been sick for over a year and received less than six months of EOL care. Tables 1 and 2 outline KII and IDI participant characteristics, respectively.

**Table 1:** Sociodemographic characteristics of key-informant interview participants

| Characteristics | Key-informant interviews (n = 20) |  |  |
| --- | --- | --- | --- |
|  | Doctors | Nurses | Total |
| **Age (years)** |  |  |  |
| 30-39 | 0 | 3 | 3 |
| 40-49 | 6 | 5 | 11 |
| 50-59 | 4 | 1 | 5 |
| 60-69 | 0 | 1 | 1 |
| **Total** | **10** | **10** | **20** |
| **Sex** |  |  |  |
| Male | 6 | 1 | 7 |
| Female | 4 | 9 | 13 |
| **Total** | **10** | **10** | **20** |
| **Marital Status** |  |  |  |
| Single | 0 | 2 | 2 |
| Married | 10 | 8 | 18 |
| Others | 0 | 0 | 0 |
| **Total** | **10** | **10** | **20** |
| **Level of education** |  |  |  |
| None | 0 | 0 | 0 |
| Primary | 0 | 0 | 0 |
| Secondary | 0 | 0 | 0 |
| Tertiary | 10 | 10 | 16 |
| **Total** | **10** | **10** | **20** |
| **Ethics training post-qualification** |  |  |  |
| Yes | 4 | 4 | 8 |
| No | 6 | 6 | 12 |
| **Total** | **10** | **10** | **20** |
| **ICU or oncology ward work duration** |  |  |  |
| < 1 year | 0 | 1 | 1 |
| 1-5 years | 1 | 2 | 3 |
| 6-10 years | 2 | 1 | 3 |
| 11-15 years | 5 | 5 | 10 |
| 16-20 years | 2 | 0 | 2 |
| > 20 years | 0 | 1 | 1 |
| **Total** | **10** | **10** | **20** |

**Table 2:**
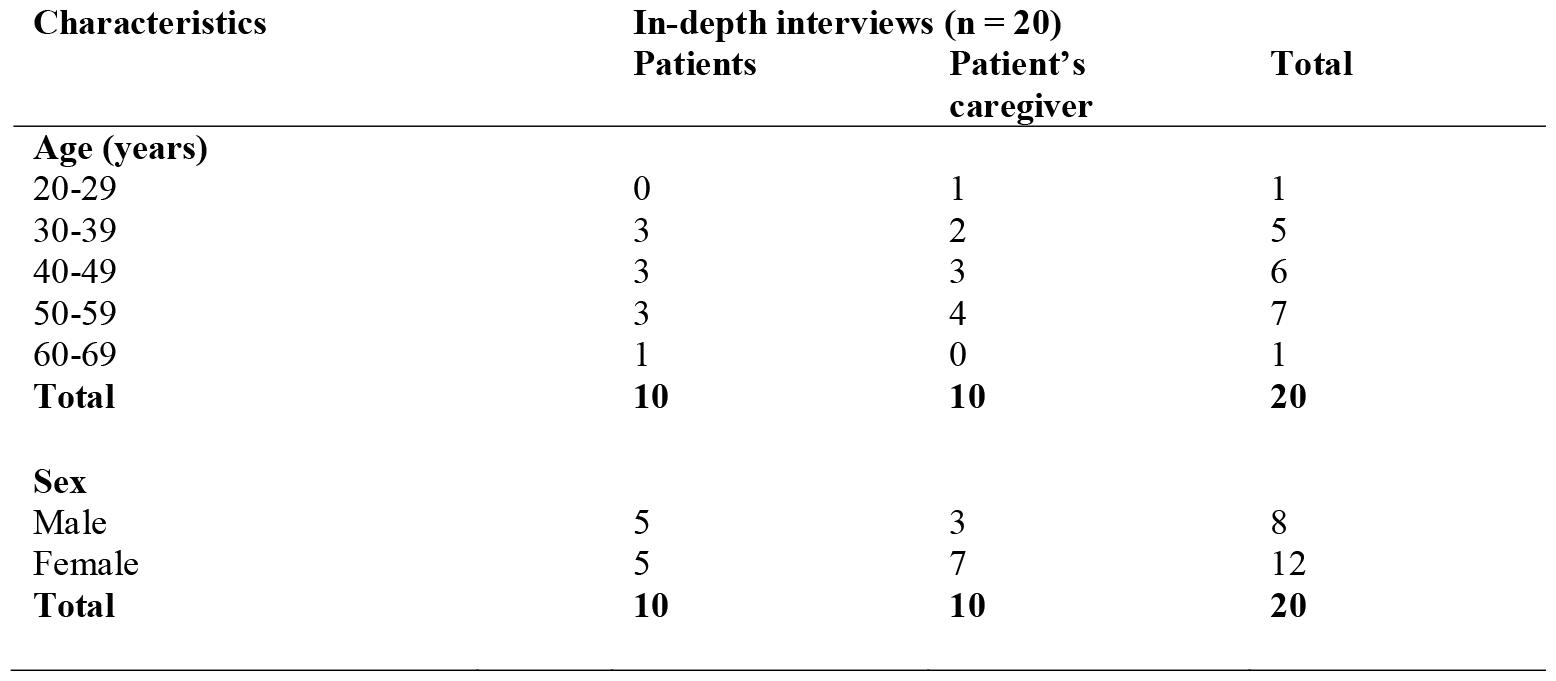
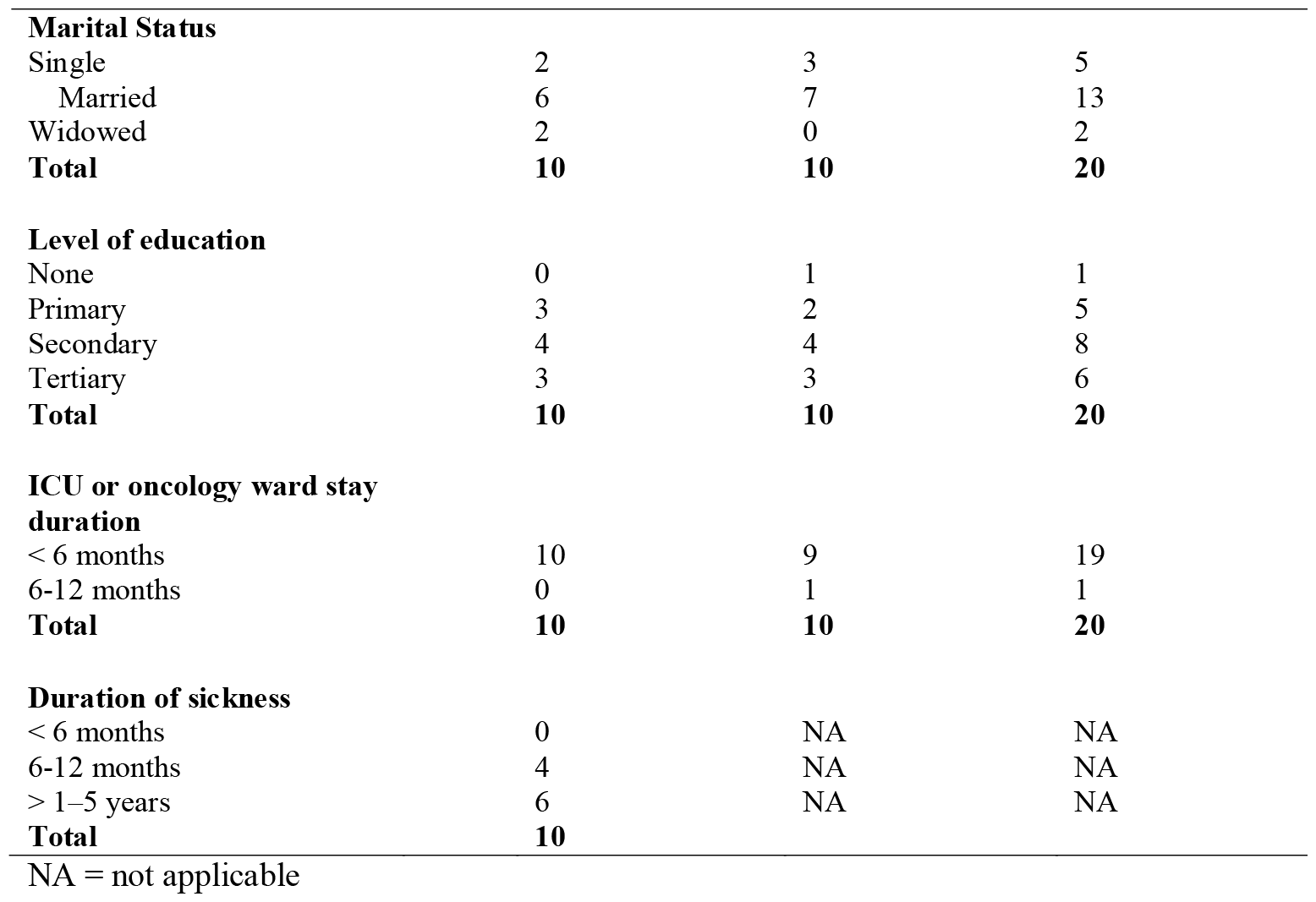
Sociodemographic characteristics of in-depth interview participants

### Occurrences of known common or usual ethical issues in EOL care

The main themes identified include incidents related to bioethical principles or their conflicts with these principles, as presented below.

### Ethical issues related to autonomy

Participants discussed medical futility, treatment refusal, truthful disclosures, family requests to withhold competent patient condition information from them, and confidentiality.

Medical futility occurs when patients request inappropriate treatment or procedures in hopes of a miraculous cure, leading some physicians to continue treatment even when clinical judgments cannot support any benefit. “*When they (the patient) are brain dead, their relatives keep insisting that one should keep oxygenating the patient”* (KII: Doctor), and *“sometimes, I yield because of their psychology; let them not feel that everybody has given up on them*.*”* (KII: Doctor). It was emphasized that *“if the patient is paying for the services, [*…*] I will not discontinue treatment*.*”* (KII: Doctor) Treatment refusal often stems from the patient religion’s beliefs. *“Like the patient who doesn’t like taking blood transfusions, […] a diabetic who could not eat fish, pork, or food prepared with these items. […] Their beliefs made it difficult in their care*.*”* (KII: Nurse) Patients often refuse treatment due to fear or ignorance. Some “*people reject oxygen when their relative gets to the point of requiring oxygen because they feel this means that their relative will die fast*.*”* (KII: Doctor). Others refuse NG tubes due to “*feeling they are not getting enough through the NG tube*.*”* (KII: Nurse). Some refuse treatment because “*they are very afraid or do not understand the disease or [*…*] things like the side effects*.*”* (KII: Doctor). A patient narrated, *“I said no to the oxygen because anyone they put the oxygen on does not usually survive*.” (IDI: patient) Truthful disclosure occurs when nurses and doctors misrepresent patients’ terminal health conditions to the patient or their family. *“They (the patients) were given false hope—don’t worry; meanwhile, they were already in the terminal phase of their health conditions*.*”* (KII: Nurse) Some worry about disclosing EOL conditions. “*The problem is [*…*] balancing the depth of information they need with their ability to understand it and not going into despair*.*”* (KII: Doctor) Another said, *“We know that these patients are going, but who will tell?”* (KII: Doctor) Some doctors face negative reactions after truthfully disclosing their patients’ conditions. *“They started […] acting as though I was a sort of obstacle for their healing*.*”* (KII: Doctor) Others believe otherwise. “*I have never experienced bad consequences from telling them the truth*.*”* (KII: Doctor) Patients and their families also face difficulties and sometimes feel deceived. *“They do not usually have time to discuss; I don’t know what is going to happen*.*”* (IDI: patient)

He commenced treatment without telling us the exact cause or nature of her illness. After 5 months, it (the breast lesion) came back again, and we went back to the same doctor only to be told that no treatment could cure her. (IDI: patient’s family caregiver)

Family members request withholding information from competent patients, posing a dilemma about *“whether to tell the patient or not*.*”* (KII: Nurse) A doctor narrated, *“I was invited to review the case […] and was cautioned that the woman remains unaware of her terminal cancer diagnosis and EOL status*.*”* (KII: Doctor) This particularly concerns *“some children; they just don’t want their elderly parents burdened with anything in their sick bed*.*”* (KII: Doctor) Health caregivers often accept withholding information, believing *“their reason is in the best interest of the patient*.*”* (KII: Doctor) A patient expressed that *“the doctors were willing to tell me about my condition, […], but my daughter would request otherwise*.*”* (IDI: patient) Some family caregivers expressed their reason: *“It may cause her more harm, and she might die off faster*.*”* (IDI: patient’s family caregiver)

Confidentiality issues arise when doctors share patient information with their families without consent. *“I may talk about the patient without their consent. Yes, in some cases*.*”* (KII: Doctor) Another shared experience:

> The wife brought him (the patient) in. I thought I could, and we conversed freely. […] I noticed that the wife was not happy. She didn’t know he had terminal cancer, and they just got married. It was after the conversations that I realized the situation. (KII: Doctor)

**Ethical issues related to beneficence and non-maleficence** include treatment limitation or withdrawal, inadequate pain management, and conflicting interests, as detailed below.

Issues of limiting or withdrawing treatment were faced by the doctors. There was *“a case of advanced cancer at EOL [*…*] and I have to withdraw all drugs. It became an issue of how to explain and carry the patient and their family along in doing this*.*”* (KII: Doctor) Patients’ relatives request treatment discontinuation for non-beneficial reasons. *“A man called our team, requesting that we remove the oxygen from the wife that he wanted her to pass on so that he could carry her and go*.*”* (KII: Doctor) Patients requesting DNR are rare, often due to pain and frustration. *“A patient requested DNR; [*…*] we put the patient on antidepressants and involved a psychologist. [*…*] The patient became happier and never mentioned it again*.*”* (KII: Doctor)

Inadequate pain management has caused significant harm to patients, leading to “*anxiety, depression, and even psychiatric manifestations*.*”* (KII: Doctor). The pain and palliative care unit is struggling with drug availability. *“For more than 3 months, there was no morphine readily available*.*”* (KII: Doctor)

> We have an irregular supply of opioids. The choice of opioids in this country is so narrow that it is either this or this. Sometimes there are lots of patients who don’t respond to option A, or the relief will not last long, or option B. (KII: Doctor)

A patient said, *“I still feel pain even though I am taking the drugs*.*”* (IDI: patient), while a relative reported: *“The patient hardly sleeps well due to pain*.*”* (IDI: patient’s family caregiver)

Inter-professional conflict arises from differing views on patient care among doctors and nurses concerning the best care options. *“A 76-80 year old breast cancer patient was advised against chemotherapy based on existing guidelines. Some doctors preferred chemotherapy despite the patient’s health status*.*”* (KII: Doctor) Conflicting instructions regarding patients also occurred: *“Doctor A will give an instruction, and then Doctor B will give contrary instructions*.*”* (KII: Doctor) and *“medical team stopping the morphine the palliative team has given occurs*.*”* (KII: Doctor)

Conflicts arise between healthcare teams and patients when patients fail to cooperate with medications and consent to procedures. *“You can give instructions about medications and treatment, but the patients will come for follow-up and tell stories*.*”* (KII: Doctor) Also, *“in consent for procedures, patients at times may be antagonistic, [*…*] want their own opinion done*.*”* (KII: Nurse) Poor communication also contributes to conflicts; *“their (health team) communication is very poor*.*”* (IDI: patient’s family caregiver)

Conflicts between patients and their families arise as interests are pursued by each party. *“Some relatives may be looking at the finances, time, and energy involved, while the patient is looking at the life he or she is going to lose*.*”* (KII: Doctor), and *“the patient does not want to go home, but his relatives want him to go home*.*”* (KII: Nurse) Conflicts in choosing between two care modalities arise due to financial implications. *“He is considering the children he is responsible for and does not want to waste money [*…*], but the relatives insist that it is life first*.*”* (KII: Doctor) *“The patient may want targeted treatment, but due to cost, the relatives may want another alternative for him*.*”* (KII: Doctor)

**Ethical issues related to justice** involve financial burdens without patient consent and healthcare system unfairness, often arising when situations are undisclosed. *“A patient was compelled to get a particular expensive treatment with an uncertain outcome [*…*] unknown to the patient or their relatives*.*”* (KII: Doctor) Also, *“the patient has stayed long in the ward. When the doctors requested brain surgery, [*…*] said, Surgery will not benefit the patient*.*”* (KII: Nurse) Inadequate care for EOL patients is often due to financial constraints. *“A patient with cancer came in bleeding; we rushed and gave her a hemostatic dose of radiotherapy; we needed to continue with the full radiotherapy, but she could not afford it*.*”* (KII: Doctor)

**An ethical issue arising from conflicting bioethics principles** is paternalism. *“A particular doctor managing EOL cases will say, Do not resuscitate my patients. [*…*] who are at the EOL but without the patient agreeing to this*.*”* (KII: Nurse) Some patients requested or considered euthanasia, but no issues were raised. *“In the height of a very critical illness or severe pain, the patient can say, I want to die; please kill me or help me to die*.*”* (KII: Doctor)

### Occurrences of uncommon or unusual ethical issues

Participants disclosed unusual ethical issues in EOL care, encompassing patients’ reluctance to discuss options, family withdrawal of support, and late presentations. It was said that *“the patients themselves would not wish to discuss terminal (EOL) issues*.*”* (KII: Doctor), and expressed that *“I prefer if I am not told*.*”* (IDI: patient, oncology-care ward) Due to stigmatization and depression, *“if they (family members) know it is a terminal case, some may withhold their support*.*”* (KII: Doctor) “*Most of them don’t even know how it started. Yet they are blamed for their problems*.*”* (KII: Nurse) Late presentations are due to delayed referrals of patients to the palliative unit. *“Most of the time, the doctor will not refer the patients on time to the palliative unit [*…*] until they enter severe distress*.*”* (KII: Nurse)

## DISCUSSION

To our knowledge, this study represents the first empirical investigation in Nigeria on ethical issues in end-of-life (EOL) care. The data presented demonstrates the breadth and depth of ethical issues encountered in EOL care in Enugu, Nigeria, and possibly sub-Saharan Africa.

The key ethical issues related to respect for patient autonomy include medical futility, treatment refusal, truthful disclosures, patient confidentiality, and family requests that the patient’s condition not be disclosed to them. Previous research in the USA,^[3][8]^ UK,^[22][23]^ Taiwan,^[16]^ Iran,^[11]^ and Portugal,^[24][25]^ found similar results, suggesting that these ethical issues may be inherent in EOL care and transcend cultural or socioeconomic situations.

We found that doctors prioritize patient hope through futile treatment, which may benefit their psychological well-being. Previous studies reported issues of medical futility. ^[3][8][11][23][24]^ It may be justifiable since overall well-being considers quality of life and psychological well-being paramount.^[26]^ However, futile treatment can be destructive to a patient’s quality of life and cause more suffering, ultimately leading to an undignified death.^[8][24][25]^ Patients must understand their EOL situation and make informed decisions.

Treatment refusals in this study were primarily verbal, stemming from fear, ignorance, or religious beliefs, contrasting with findings in developed countries where advance directives are common.^[7][26]^ Refusals due to religious beliefs are not limited to EOL care,^[8]^ and families refusing treatments on behalf of the patients are reported.^[3][27]^ Treatment withdrawal issues were found in our study when patients and families do so for reasons beyond risk-benefit analyses. Similar issues occur in the US^[13]^ and Scotland,^[31]^ suggesting the need for proactive guidelines, such as advance directives, to address withdrawal concerns.

Health caregivers’ concerns and cultural perceptions hinder truthful disclosure, with educational status influencing doctors’ choices in this study. Studies in USA,^[3][8]^ Iran,^[11]^ Taiwan,^[16]^ the UK,^[22]^ and Portugal,^[24]^ have reported issues of truthful disclosure. Lack of formal training in communication skills contributes to this issue.^[11]^ However, culturally approved manners are suggested for the most truthful disclosure.^[28][23]^ This highlights the need for training and regular updates for healthcare providers to communicate effectively. Our study also found that family members request that the clinical details of a competent patient receiving EOL care be withheld from them, believing it is in their best interests. Similar findings exist in reported studies.^[3][11][16]^ While family involvement is acknowledged,^[14][16]^ a low “good death score” is observed among those unaware of imminent death,^[29]^ suggesting undisclosed circumstances may cause harm.

Confidentiality issues were described in this study, with some patients concerned about sharing information with family members at the EOL without their consent, which occurs due to the belief that individuals don’t mind if it is shared with their family. A similar study discovered that this issue is common when family members request exclusive information about a patient’s condition.^[30]^ Our study found that, despite good intentions, harm to the patient may be inevitable if confidentiality is violated.

Participants in this study experienced limited pain control issues due to the inadequate supply and availability of pain-relieving drugs. Patients in India reported pain relief accessibility issues.^[10]^ This could be because it is a regulated class of medication. Similar to a report in America,^[3]^ this study highlights instances where doctors discontinue pain-relieving drugs prescribed by the pain and palliative care unit. The need for education and re-education of caregivers and policymakers on the current pain management protocol is crucial for improving available and proper pain control.

The inter-professional conflict discovered in our study is similar to that previously reported in the USA,^[3][32]^ and could be attributed to differing patient care perspectives and communication gaps. While professional hierarchy could explain this finding,^[23]^ nurses’ increased involvement in patient care decisions necessitates collaboration,^[33]^ which necessitates conflict anticipation and mitigation.

Funds limited the access of patients to essential care in our study. However, in Iran, the high cost of similar services did not prevent physicians from choosing standard treatment for well-informed patients.^[11]^ An organizational barrier could explain our research findings, as the Nigerian healthcare insurance system does not cover EOL care. Advocacy for expanded coverage is needed.

This study uncovered rare ethical issues in EOL care, such as patients’ reluctance to discuss death, family withdrawal due to stigma, and blaming patients for their condition. Families play a crucial role in EOL care, often providing support and decision-making.^[38][39][40]^ Lack of preparation and support may contribute to their struggles. The findings underscore the need for better support for family caregivers. Although late-onset palliative care is common, it’s seldom discussed as an ethical issue, as found in our study,^[41]^ which emphasizes the importance of timely diagnosis and referral to prevent harm. Training caregivers to deliver difficult news could enhance patient referrals.^[41]^

### Strengths of this study

Our study includes multiple participants and provides diverse perspectives and a better understanding of ethical issues in EOL care. Also, a classification based on the infringed bioethics principles to address ethical issues in EOL care was used, allowing for comprehensive reporting and analysis of this issue in a way not previously considered. This classification could be used globally.

### Limitation/recommendation

Health caregivers in EOL care should have the knowledge and skills necessary to engage patients and their families in dialogue on EOL issues. It was not explored in this study. Proactive guidelines such as advance directives should be explored and adopted to fit the local peculiarities of the study area.

## CONCLUSION

EOL care at the UNTH-E oncology center and ICU raises ethical concerns for patients, families, and medical staff. This study identifies common ethical issues in EOL care in this part of Nigeria, including violations of patients’s autonomy linked to medical futility, truthful disclosure, family requests that a competent patient not be informed about his condition, confidentiality; issues of beneficence and nonmaleficence such as limiting or withdrawing a treatment, limited or insufficient pain management, conflicting interests between concerned parties; issues of justice such as financial burdens imposed on patients without their consent; and conflicting bioethical principles such as paternalism. Furthermore, there were occurrences of some uncommon or unusual types of ethical issues in EOL care, such as patients refusing to be told that they are now receiving EOL care or discussing issues about death or dying, withdrawal of family support and stigmatization, late presentation, early detection, or delayed referrals.

## Data Availability

All data produced in the present study are available upon reasonable request to the authors

